# Early transmission of SARS-CoV-2 in South Africa: An epidemiological and phylogenetic report

**DOI:** 10.1101/2020.05.29.20116376

**Authors:** Jennifer Giandhari, Sureshnee Pillay, Eduan Wilkinson, Houriiyah Tegally, Ilya Sinayskiy, Maria Schuld, Jose Lourenco, Benjamin Chimukangara, Richard Lessells, Yunus Moosa, Inbal Gazy, Maryam Fish, Lavanya Singh, Khulekani Sedwell Khanyile, Vagner Fonseca, Marta Giovanetti, Luiz Carols Alcantara, Francesco Petruccione, Tulio de Oliveira

## Abstract

**Background:** The emergence of a novel coronavirus, SARS-CoV-2, in December 2019, progressed to become a world pandemic in a few months and reached South Africa at the beginning of March. To investigate introduction and understand the early transmission dynamics of the virus, we formed the South African Network for Genomics Surveillance of COVID (SANGS_COVID), a network of ten government and university laboratories. Here, we present the first results of this effort, which is a molecular epidemiological study of the first twenty-one SARS-CoV-2 whole genomes sampled in the first port of entry, KwaZulu-Natal (KZN), during the first month of the epidemic. By combining this with calculations of the effective reproduction number (R), we aim to shed light on the patterns of infections that define the epidemic in South Africa.

**Methods:** R was calculated using positive cases and deaths from reports provided by the four major provinces. Molecular epidemiology investigation involved sequencing viral genomes from patients in KZN using ARCTIC protocols and assembling whole genomes using meticulous alignment methods. Phylogenetic analysis was performed using maximum likelihood (ML) and Bayesian trees, lineage classification and molecular clock calculations.

**Findings:** The epidemic in South Africa has been very heterogeneous. Two of the largest provinces, Gauteng, home of the two large metropolis Johannesburg and Pretoria, and KwaZulu-Natal, home of the third largest city in the country Durban, had a slow growth rate on the number of detected cases. Whereas, Western Cape, home of Cape Town, and the Eastern Cape provinces the epidemic is spreading fast. Our estimates of transmission potential for South Africa suggest a decreasing transmission potential towards R=1 since the first cases and deaths have been reported. However, between 06 May and 18 May 2020, we estimate that R was on average 1.39 (1.04–2.15, 95% CI). We also demonstrate that early transmission in KZN, and most probably in all main regions of SA, was associated with multiple international introductions and dominated by lineages B1 and B. The study also provides evidence for locally acquired infections in a hospital in Durban within the first month of the epidemic, which inflated early mortality in KZN.

**Interpretation:** This first report of SANGS_COVID consortium focuses on understanding the epidemic heterogeneity and introduction of SARS-CoV-2 strains in the first month of the epidemic in South Africa. The early introduction of SARS-CoV-2 in KZN included caused a localized outbreak in a hospital, provides potential explanations for the initially high death rates in the province. The current high rate of transmission of COVID-19 in the Western Cape and Eastern Cape highlights the crucial need to strength local genomic surveillance in South Africa.

**Funding:** UKZN Flagship Program entitled: Afrocentric Precision Approach to Control Health Epidemic, by a research Flagship grant from the South African Medical Research Council (MRC-RFA-UFSP-01- 2013/UKZN HIVEPI, by the the Technology Innovation Agency and the the Department of Science and Innovation and by National Human Genome Re- search Institute of the National Institutes of Health under Award Number U24HG006941. H3ABioNet is an initiative of the Human Health and Heredity in Africa Consortium (H3Africa).

**Research in context Evidence before this study:** We searched PubMed, BioRxiv and MedRxiv for reports on epidemiology and phylogenetic analysis using whole genome sequencing (WGS) of SARS-CoV-2. We used the following keywords: SARS-CoV-2, COVID-19, 2019-nCoV or novel coronavirus and transmission genomics, epidemiology, phylogenetic or reproduction number. Our search identified an important lack of molecular epidemiology studies in the southern hemisphere, with only a few reports from Latin America and one in Africa. In other early transmission reports on SARS-CoV-2 infections in Africa, authors focused on transmission dynamics, but molecular and phylogenetic methods were missing.

**Added value of this study:** With a growing sampling bias in the study of transmission genomics of the SARS-CoV-2 pandemic, it is important for us to report high-quality whole genome sequencing (WGS) of local SARS-CoV-2 samples and in-depth phylogenetic analyses of the first month of infection in South-Africa. In our molecular epidemiological investigation, we identify the early transmission routes of the infection in the KZN and report thirteen distinct introductions from many locations and a cluster of localized transmission linked to a healthcare setting that caused most of the initial deaths in South Africa. Furthermore, we formed a national consortium in South Africa, funded by the Department of Science and Innovation and the South African Medical Research Council, to capacitate ten local laboratories to produce and analyse SARS-CoV-2 data in near real time.

**Implications of all the available evidence:** The COVID-19 pandemic is progressing around the world and in Africa. Early transmission genomics and dynamics of SARS-CoV-2 throw light on the early stages of the epidemic in a given region. This facilitates the investigation of localized outbreaks and serves to inform public health responses in South Africa.

## Introduction

The novel coronavirus disease 2019 (COVID-19) was detected in China in late December 2019. On 30 January 2020, it was declared a Public Health Emergency of International Concern by the World Health Organization (WHO) (1). By 15 May 2020, there were 4,621,410 COVID-19 cases and 308,542 related deaths (2) worldwide involving almost every country in the world. Within five months, the virus had spread to Europe, America and eventually to Africa. The first case in Africa was reported in Nigeria on 28 February 2020 (3), and at the time of writing, the pandemic has spread to almost all countries on the African continent. South Africa has had the highest number of COVID-19 cases to date with a total of 13,524 people infected and 247 deaths (as at 15^th^ May)(4).

The first confirmed case of COVID-19 in South Africa was reported on 5 March 2020. Decisive early action was taken by the government: a national state of disaster was declared on 15 March 2020, and a nationwide lockdown was enforced on 27 March 2020 to avoid the first wave overwhelming the health system. While initially only people who had travelled to at-risk countries and their contacts received PCR tests for severe acute respiratory syndrome-related coronavirus 2 (SARS-CoV-2), the recommendation broadened to include all people with an acute respiratory illness. Furthermore, a program of community-based screening and testing was rolled out across the country (4). Testing increased rapidly and by the middle of May 2020, over 600,000 tests had been carried out in South Africa (approximately 10,000 per million population) (5).

As the global pandemic has expanded, WGS and genomic epidemiology (6) have been consistently used to investigate COVID-19 transmission and outbreaks (7–13). In response to the COVID-19 pandemic, the South African Network for Genomics Surveillance of COVID (SANGS_COVID) was formed, which is a network of five large government laboratories and five public universities funded by the Department of Science and Innovation and the South African Medical Research Council. In this paper, our consortium focuses on a detailed analysis of the epidemic in South Africa and preliminary genomic analysis of some of the first introductions of SARS-CoV-2 in KZN. We show that although the South African epidemic started in KZN, which have the first cases and deaths, other provinces in the country, namely the Western Cape (WC), Gauteng (GP) and the Eastern Cape (EC), have overtaken KZN in the number of confirmed cases. We also show evidence of many distinct introductions of SARS-CoV-2 in KZN and early evidence suggesting nosocomial transmission.

## Results

### Epidemiology of COVID-19 in KZN and South Africa

The first confirmed case of COVID-19 in South Africa was reported on 5 March 2020 in KZN, a South African citizen returning home from a skiing holiday in Italy. A steady increase in the number of confirmed cases in South Africa (all imported cases) followed over the next week, with the first suspected case of local transmission reported on 13 March 2020 in Durban, KwaZulu-Natal. The early cases were predominantly located in the three provinces with the main urban populations and international travel hubs, namely GP (main cities Pretoria and Johannesburg), the WC (Cape Town) and KZN (Durban).In these three provinces, the doubling time for confirmed cases was approximately three days prior to the lockdown (Figure 1). However, since the lockdown on 27 March 2020, the epidemic seems to be growing at different rates in South Africa.

**Figure 1:**
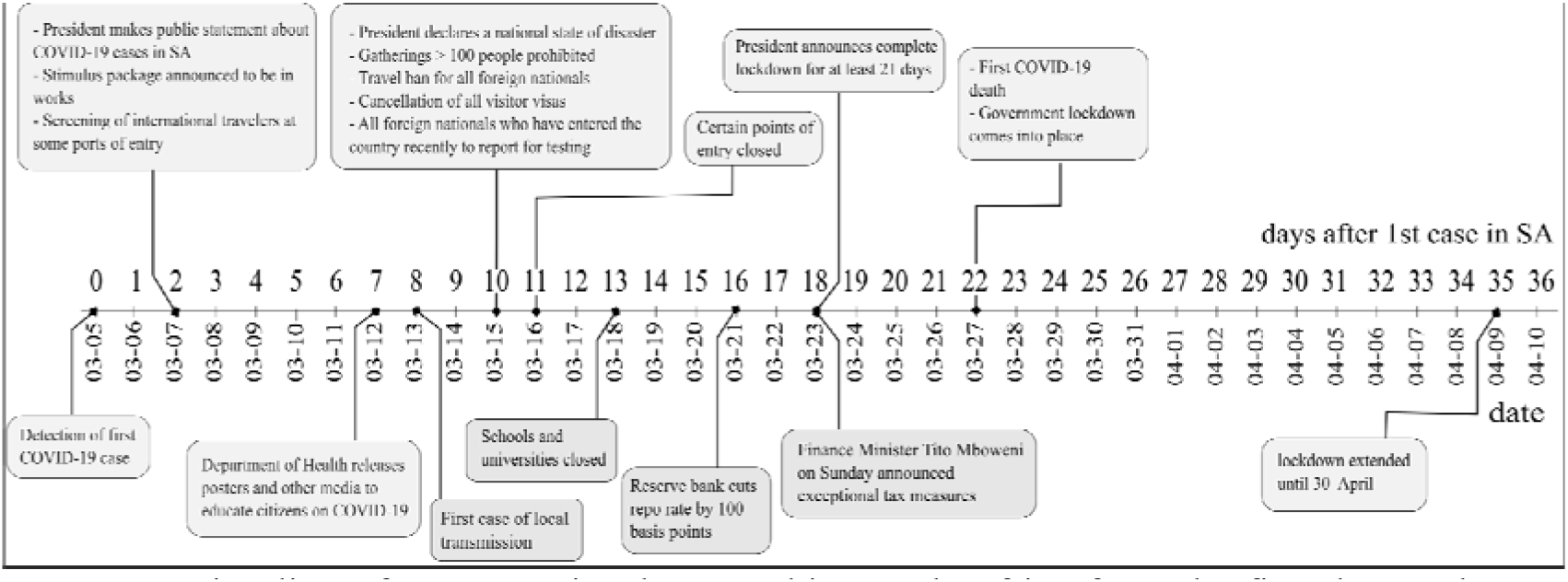
Timeline of measures implemented in South Africa from the first detected COVID-19 case on 5 March to the expansion of the lockdown in April 2020.

The South African epidemic has been very heterogeneous. For example, the first cases and deaths happened in KZN and GP. This was more pronounced in KZN, as a large nosocomial outbreak in a private hospital in Durban caused KZN to lead the country in number of deaths until the WC overtook it on 21 April 2020. In addition, GP, home of the largest metropolitan area of Johannesburg, had an unusual epidemic, as the majority of initial cases were in middle age and rich individuals who traveled overseas for holidays. This translated in a very small number of deaths over time and infections were concentrated in the rich suburb of Sandton in Johannesburg. However, the epidemic seems to be expanding the fastest in the Western Cape, specially in Cape Town. At the time of writing this report, this province has over 60% of all of the cases and deaths in South Africa (Figure 2). There is mounting evidence that the Western Cape is seeding the growing epidemic in the Eastern Cape as the funerals from some of the deaths in the Western Cape is taking place in the Eastern Cape (REF).

**Figure 2:**
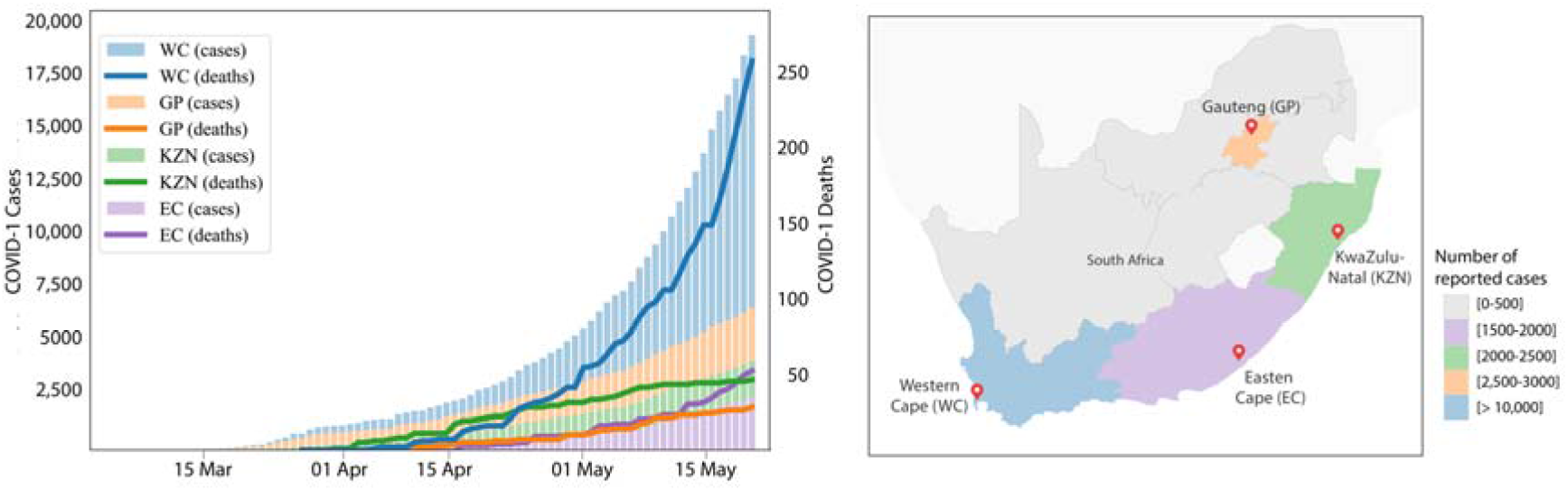
Summary of the COVID-19 epidemic in South Africa. A) Numbers of COVID-19 cases and deaths in the Western Cape (WC), Gauteng (GP), KwaZulu-Natal (KZN) and the Eastern Cape (EC). B) Geographic map showing the location of South African provinces.

This dynamic and heterogeneous epidemic complicates the estimation of effective reproductive number (R) over time. For example, deaths, which is normally one of the gold standard data for estimation of R0 for South Africa in May 2020 has been stable at 1·12 (1.0–1.2) (Supplementary figure 1). Our estimates of transmission potential for South Africa suggest a decreasing transmission potential towards R=1 since the first cases and deaths have been reported. However, by the last period analyzed between 06-May and 18-May, using the Wallinga et al approach, we estimate that R was on average 1.39 (1.04–2.15, 95% CI). Given the low deaths from the country and the heterogeneous distribution, deaths seem to not to allow the estimation of R0 from individual provinces. For example, KZN, the first province affected by COVID-19, initially had the highest death rate but in the last period analyzed, had only 3 deaths. Given that deaths do not seen to be effective for R estimation in the different provinces, so attempted to estimate R from COVID-19 positive cases (Supplementary Figure 2, Supplementary Table S1).

### SARS-CoV2 genomes from KZN

In order to determine the route of introduction of the SARS-CoV-2 in KZN, we assessed 27 of some of the first confirmed cases in the province. Samples obtained from nasopharyngeal swabs represented fourteen females and ten males between the ages of 23–74 years. We managed to produce 20 near-whole genome sequences (>90% coverage) from these samples, and six partial genomes (Supplementary Table S2, Table S3). To this dataset, we added an extra genome from the NICD, which was sampled in KZN (a close contact of the first reported case) on 7 March 2020. The 21 KZN whole genomes (20 KRISP and one NICD) were assigned to SARS-CoV-2 sub-lineages according to the nomenclature proposed and lineage classification obtained from > 5000 genomes analyzed by Rambaut et al. (14). Given uncertainties pertaining to the low diversity of this virus (15), we restricted lineage assignment to the four most prominent subgroups (A, B, B·1 and B·2). Of the 21 KZN isolates being investigated in the present study, one was assigned to lineage B (KRISP-006) and one to sub-lineage B·2 (KRISP-002) (Figure 3). The remaining 19 KZN sequences were all assigned to lineage B·1. The B·1 lineage consists primarily of cases originating in Europe (Figure 3c), suggesting that introductions from Europe accounted for many of the early cases in KZN.

**Figure 3:**
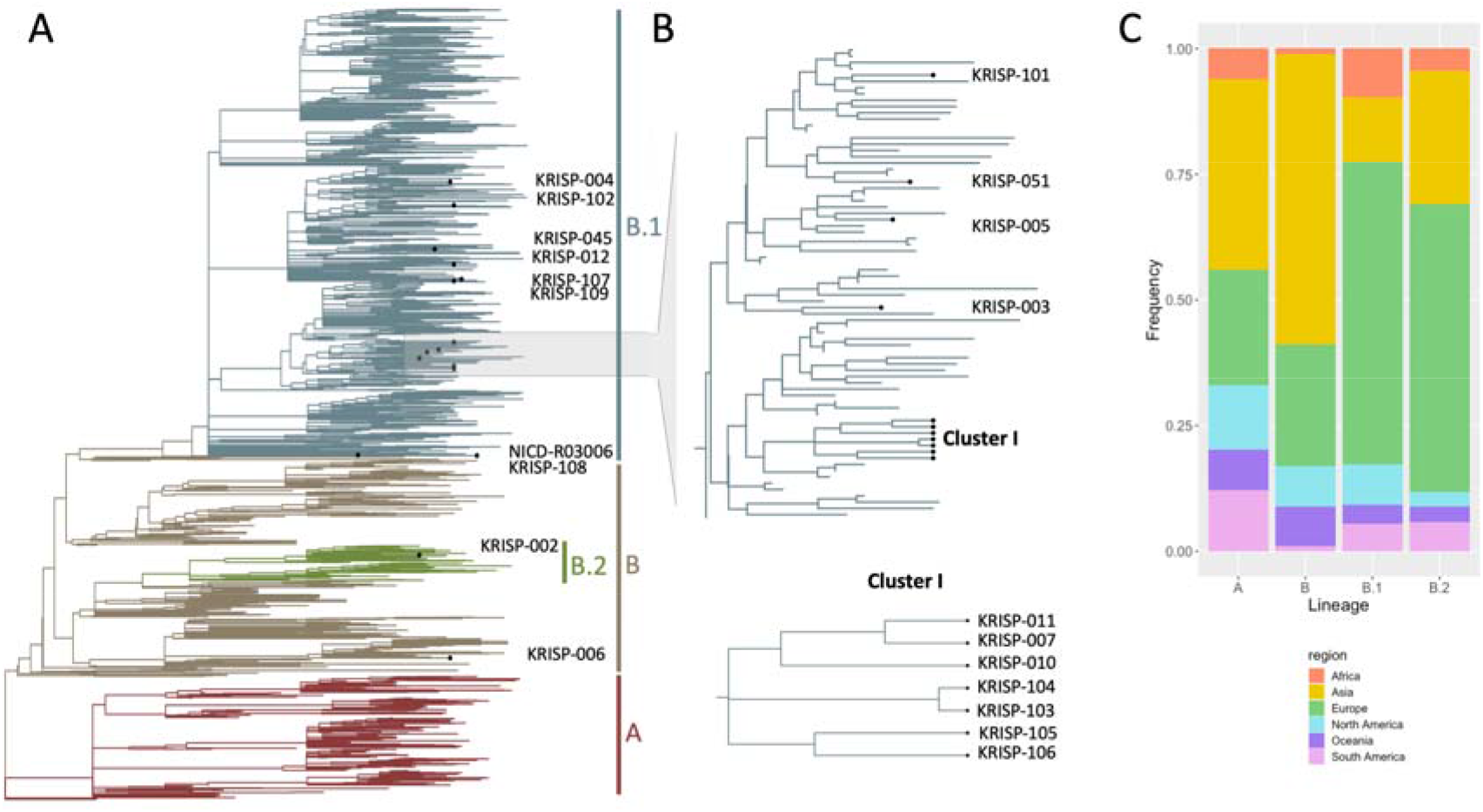
Phylogenetic analysis. (A) A time scaled Maximum likelihood tree of 1849 sequences including 21 genotypes from KwaZulu-Natal, South Africa. Major lineages of SARS-CoV-2 are labelled. B) Monophyletic cluster of KZN sequences. (C) Stacked barplot showing the lineage breakdown of the dataset by region.

Although our investigation contained only a small number of samples from the first month of the epidemic in South Africa, we identified at least 13 distinct introductions (Figure 3 and Figure 4) and one monophyletic cluster involving seven sequences. Three of the sequences (*KRISP-007, KRISP-010* and *KRISP-011*) were identical and contained five mutations (241C>T, 3037C>T, 14408C>T, 16376C>T and 23403A>G). After investigation, we found that these samples were from health care workers at a private hospital in Durban, KZN, with no history of travel outside the country. A detailed investigation is currently being conducted in this hospital, but preliminary findings suggest a point-source nosocomial outbreak (Lessells et al. manuscript in preparation). The other four sequences in the cluster contained two pairs (*KRISP-103; KRISP-104* and *KRISP-105; KRISP-106*). Samples 103 and 104 are identical to one another and are characterized by three additional mutations (5672C>A, 10592A>G, and 26063G>T) on top of the ones reported above (Supplementary Table S4). Samples 106 has acquired one additional mutation (24034C>T) on top of the five mutations common to the hospital outbreak, while sample 105 acquired another two mutation (13766A>T and 18411T>C) on top of the mutations found in 106. These two pairs were derived from random sampling within the Durban metropolitan area suggesting early evidence of localized transmission in Durban (Figure 3b).

### Time-resolved analysis of three main lineages circulating in KZN

To determine the evolutionary relationship of the KZN sequences to the world-wide SARSCoV-2 pandemic, we conducted a Bayesian molecular clock analysis for each of the lineages found in KZN (Figure 4). Coalescent molecular clock analyses of lineage B places KRISp-006 at the base of a subclade along with a sequences from Canada with high posterior support (*P*=1.0). The remainder of the subclade contains sequences from a large number of Asia countries (Singapore, Phillippines & Malaysia), Australia, the United States and the United Kingdom. The B·1 Bayesian analysis, which contained 19 KZN sequences, suggest multiple introductions into KZN from European countries. Due to low diversity in this lineage the posterior support for splits in the tree were very low. Furthermore, due to the small number of nucleotide differences between isolates the monophyletic clade of seven KZN sequences observed previously now only contained five sequences. Samples 103 and 104, though clustering together with one another, were seperated from the rest of the clade by other European reference sequences. The time to the most recent common ancestor (tMRCA) for the monophyletic KZN clade of five sequences were inferred around 23 March 2020, with the 95% Highest Posterior Density between 10 and 31 March 2020, which is consistent with the dates of the nosocomial outbreak in Durban (Lessells et al. manuscript in preparation).

**Figure 4:**
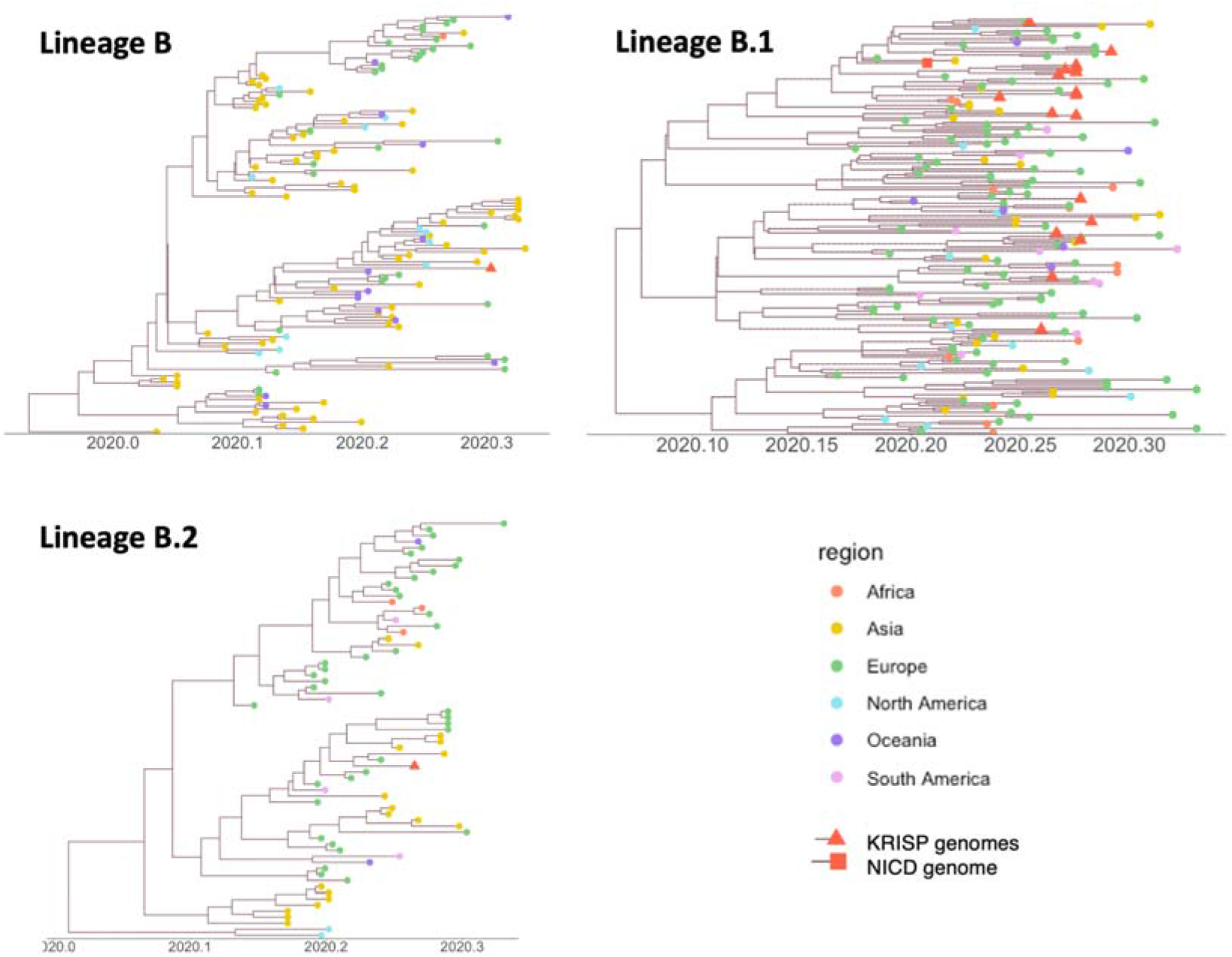
Time stamped phylogenetic trees of the three lineages of SARS-CoV-2 found in KwaZulu-Natal (KZN). The genomes produced in this study are marked with a red triangle, and the NICD genome by a red square. The geographic region of the other sequences is marked with coloured circles.

Our coalescent analyses in BEAST placed the origin of B & B·1 around the first week of December 2019 [95% HPD], with the 95% highest posterior density (HPD) ranging between mid October and the last week of December 2019. Coalescent analyses placed the time to the most recent common ancestor (MRCA) for sub-lineage B·2 around late December 2019 (95% HPD mid November 2019 – first week of January 2020). Our temporal estimates are consistent with the SARS-CoV-2 date of origin.

## Discussion & Conclusion

The spread of SARS-CoV-2 across the globe has given rise to one of the largest evolving pandemics in modern times. South Africa currently has the highest number of infections in Africa. South Africa seems to be moving to the next stage of the COVID-19 pandemic, with increasing community transmission even during the stringent lockdown and the epidemic growing at different rates in different regions of the country. At the time of writing this report, Cape Town, the main city in the WC, has the fastest increase (of new) of new infections and deaths in South Africa. Recent data indicates that over 62% of the new infections and deaths are happening in this province, although only 17% of the South African population lives in this region. The fast spread of COVID-19 in the WC is not fully explained by the higher testing rates as this province has performed between 20–22% of the tests in South Africa, but the positivity rate has been around 9%, were as in the other provinces the positivity rate is around 1–2% (ref NICD). Our estimates of transmission potential for South Africa suggest a decreasing transmission potential towards R=1 since the first cases and deaths have been reported. By last period analyzed between 06-May and 18-May, when using the Wallinga et al estimation approach applied to time series of reported cases, we estimate that R was on average 1.39 (1.04–2.15, 95% CI). Overall, these results suggest of an epidemic still in expansion at that time, in spite of a very early lockdown.

Sequencing of viral isolates from early COVID-19 cases in KZN, which is the province of South Africa with the first infections and early deaths, provided useful insights into the origins and transmission of SARS-CoV-2. From the first twenty-one genomes analyzed, we found thirteen independent introductions in KZN. These introductions were related to lineages B, B·1 and B·2, which have spread widely in Europe and North America. We also found a cluster of cases in health care workers in Durban, highlighting the potential importance of nosocomial transmission in this pandemic and potentially two other small transmission pairs. The production of genomes from the WC will be crucial to understand the drivers of transmission during the lockdown period, and particularly whether health care facilities, prisons, workplaces and other institutions are acting as amplifiers of transmission. This is one of the main activities that our consortium, SANGS-COVID, is currently working on.

Genomic analysis of SARS-CoV-2 in Africa has proved challenging on many fronts. First, sequencing of high-quality SARS-CoV-2 genomes is not a straightforward task. For example, a survey of thousands of sequences deposited in public databases has revealed a number of putative sequencing issues that appear to be the result of contamination, recurrent sequencing errors or hypermutability (16). These might arise from laboratory-specific techniques of sample preparation, sequencing technology or consensus calling. Furthermore, the low diversity of this virus and the small number of mutations that define lineages have prompted caution in the interpretation of early phylogenetic analysis worldwide (13). Often apparent local transmission clusters can in fact be the result of multiple introductions from under-sampled regions from non-uniform sequencing efforts (6,17). To mitigate this we confirmed phylogenetic results by manual inspection of mutations relative to the reference of SARSCoV-2 (Supplementary Table S4). Second, the pandemic is still evolving and grouping of SARS-CoV-2 into lineages and sub-clades is likely to be dynamic at this stage and it is influenced by proportionally larger number of sequences produced in the northern hemisphere (14). Third, the travel histories of apparent community transmission need to be thoroughly investigated in order to elucidate the true dynamics of transmission in a particular area. In our case, a subsequent investigation into the samples comprising the monophyletic cluster revealed the association with a big hospital outbreak of SARS-CoV-2 infections in Durban, KZN (Lessells et al. manuscript in preparation 2020).

This paper has some important limitations. The first is related to estimation of R0 from COVID-19 a limited number of deaths in a heterogeneous epidemic. The second is a lack of well set up genomics laboratories that can sequence the virus in Africa. This is also amplified by the difficulty of acquiring reagents that are in high demand, coupled with the disruption of air freight. It is therefore a high priority for our consortium, SANGS-COVID, to evaluate and share protocols among national laboratories in South Africa that could generate sequences of high-quality and capacitate our laboratories with the protocols and bioinformatics pipelines to properly investigate virus introduction and to validate the call of variants with a detailed and reliable bioinformatics system. SANGS-COVID is also working with the Africa Center for Disease Control (CDC) and the World Health Organization (WHO) to strengthen genomics surveillance in the African continent.

In this paper, we provide an early analysis of COVID-19 pandemic in South Africa, showing very heterogeneuos epidemics in the different provinces. We also estiamted SARS-CoV-2 genetic diversity in KZN using the first twenty one genomes from some of the first cases in the country. We find that KZN had many distinct introductions of SARS-CoV-2, but also had early evidence of nosocomial transmission. The pandemic at the local level is still developing and the objective of SANGS-COVID is to clarify the dynamics of the epidemic in South Africa and devise the most effective measures as the outbreak evolves.

## Methods

### Data sources

We used publicly released data up to 31 March 2020 from the National Department of Health (NDoH) and the NICD in South Africa, which (are) is collected in the repository of the Data Science for Social Impact Research Group at the University of Pretoria (18), as well as global data on confirmed cases from the Johns Hopkins Coronavirus Resource Centre (19). The NDoH releases daily updates on the number of new confirmed cases, with a breakdown by province. In the early stages of the epidemic, individual-level information on sex, age and travel history was released, but detailed reporting was discontinued on 23 March. In addition, the National Institute of Communicable Diseases (NICD) releases daily updates on the number of reverse-transcriptase polymerase chain reaction (RT-PCR) tests performed across all public and private sector laboratories, as well as the number of cases testing positive for severe acute respiratory syndrome-related coronavirus 2 (c). We also extracted information from government press releases and speech transcripts to chart a timeline of the government response to the epidemic. To understand the epidemic trajectory, we plotted the cumulative number of confirmed cases by province since the report of the hundredth case in the country by province.

### Epidemiological analysis and reproductive number estimation

The effective reproduction number (R) was estimated by taking into account the observed epidemic growth rate r and two theoretical relationships (i, ii) of R with r previously described in the literature. (i) We used the relationship R=(1+r/b)^a^ as described in Imperial College London’s COVID-19 report 13 (20), where a=m^2^/s^2^ and b=m/s^2^, m the serial interval (SID) mean and s the SID standard deviation. The SID distribution used is the one estimated by Nishiura and colleagues (21), with m=4.7 and s=2.9. We term this approach the Flaxman et al. approach. (ii) We used the relationship R = (1+r/sigma)(1+r/delta), with 1/sigma the infectious period and 1/delta the incubation period, as described by Wallinga and Lipsitch (22), which is based on an SEIR modelling framework and expects both periods to be exponentially distributed. We used exponential distributions with mean 5.1 days for incubation (23,24) and 4 days for the infection (23,24). We term this approach the Wallinga et al. approach. To obtain the epidemic growth rate r, we used maximum likelihood estimation in R (function optim), by fitting the exponential growth model A_0_e^rt^ to the reported time series of cases and deaths (independently), where t is time,_A0_ is the number of reports at t=0, and r the growth rate.

### Ethics statement

The project was approved by University of KwaZulu-Natal Biomedical Research Ethics Committee. Protocol reference number: BREC/00001195/2020. Project title: COVID-19 transmission and natural history in KwaZulu-Natal, South Africa: Epidemiological Investigation to Guide Prevention and Clinical Care.

### SARS-CoV-2 sample collection and preparation

Remnant samples from nasopharyngeal and oropharyngeal swab sample or extracted RNA. The swab samples were heat inactivated in a water bath at 60°C for 30 minutes, in biosafety level 3 laboratory, prior to RNA extraction. RNA was extracted using the Viral NA/gDNA Kit on the Chemagic 360 system (Perkin Elmer, Hamburg, Germany) using the automated Chemagic 360 insturment (Perkin Elmer, Hamburg, Germany) or manually using the Qiagen Viral RNA Mini Kit (QIAGEN,California, USA).

### Real Time RT-PCR

In order to detect the SARS-CoV-2 virus by PCR, the TaqPath COVID-19 CE-IVD RT-PCR Kit (Life Technologies, Carlsbad, CA) was used according to the manufacturer’s instructions. The assays target genomic regions (ORF1ab, S protein and N protein) of the SARS-CoV-2 genome. RT-PCR was performed on a QuantStudio 7 Flex Real-Time PCR instrument (Life Technologies, Carlsbad, CA). Cycle thresholds (Ct) was analysed using auto-analysis settings with the threshold lines falling within the exponential phase of the fluorescence curves and above any background signal. To accept the results, we confirmed a Ct value for RNAse P (i.e. an endogenous internal amplification control) and or the target gene in each reaction, with undetermined Ct values in the no template control. The Ct values were reported for each target gene.

### Tiling Polymerase Chain Reaction

cDNA synthesis was performed on the RNA using random primers followed by gene specific multiplex PCR using the ARTIC protocol (25). Briefly, extracted RNA was converted to cDNA using the Protoscript II First Strand cDNA synthesis Kit (New England Biolabs, Hitchin, UK) and random hexamer primers. SARS-CoV-2 whole genome amplification by multiplex PCR was carried out using primers designed on Primal Scheme (http://primal.zibraproject.org/) to generate 400bp amplicons with an overlap of 70bp that Remnant samples from nasopharyngeal and oropharyngeal swabs collected from symptomatic patients, were used for SARS-CoV-2 WGS. These samples comprised of either California, USA). above any background signal. To accept the results, we confirmed a Ct value for RNAse P covers the 30Kb SARS-CoV-2 genome. PCR products were cleaned up using AmpureXP purification beads (Beckman Coulter, High Wycombe, UK) and quantified using the Qubit dsDNA High Sensitivity assay on the Qubit 4.0 instrument (Life Technologies Carlsbad, CA).

### Illumina MiSeq Sequencing

PCR products for samples yielding sufficient material were included in this sequencing platform. Illumina® TruSeq® Nano DNA Library Prep kits were used according to the manufacturer’s protocol to prepare uniquely indexed paired end libraries of genomic DNA. The libraries were quantified using the Qubit dsDNA High Sensitivity assay on the Qubit 4.0 instrument (Life Technologies) and the fragments were analysed using the LabChip GX Touch (Perkin Elmer, Hamburg, Germany). Sequencing libraries were normalized to 4nM, pooled and denatured with 0·2N sodium acetate. 12pM sample library was spiked with 1% PhiX (PhiX Control v3 adapter-ligated library used as a control). Libraries consisting of 12 samples each were loaded onto a 500-cycle MiSeq Nano Reagent Kit v2 nano v2 Miseq reagent kit and run on the Illumina MiSeq instrument (Illumina, San Diego, CA, USA).

### Bioinformatics assembly of genomes

Raw reads coming from both Nanopore and Illumina sequencing were assembled using Genome Detective 1·126 (https://www.genomedetective.com/) and the Coronavirus Typing Tool (26, 27). The initial assembly obtained from Genome Detective was polished by aligning mapped reads to the references and filtering out low-quality mutations using bcftools 1·7–2 mpileup method. All mutations were confirmed visually with bam files using Geneious software (Biomatters Ltd, New Zealand). All of the sequences were deposited in GISAID (https://www.gisaid.org/)(23).

### Reference dataset

We downloaded all sequences and associated metadata from the GISAID sequence database (https://www.gisaid.org/) (28) as of 1 May 2020 (n=15,793). Due to the low variability of SARS-CoV-2, we wished to only include high quality sequences in our downstream analyses. To this end, we filtered out sequences that were < 25kbp in length as well as sequences with a high proportion of ambiguous sites (>5%). Additionally, we also removed sequences that lacked any geographic and or sampling date information. The resulting 10,959 sequences were analyzed along with 20 sequences that were generated by the laboratory at the KwaZulu-Natal Research Innovation and Sequencing Platform (KRISP). The dataset also contained one additional KZN sequences (*EPI_ISL_417186*) that were generated by the National Institute for Communicable Diseases (NICD) and represent a distant contact of the first diagnosed case in South Africa.

### Lineage classification

Currently, no established nomenclature system exists for SARS-CoV-2. A dynamic lineage classification method proposed by Rambault et al., was used in this study (14) via the Phylogenetic Assignment of named Global Outbreak LINeages (PANGOLIN) software suite (https://github.com/hCoV-2019/pangolin). This is aimed at identifying the most samples each were loaded onto a 500-cycle MiSeq Nano Reagent Kit v2 nano v2 Miseq reagent kit and run on the Illumina MiSeq instrument (Illumina, San Diego, CA, USA). epidemiologically important lineages of SARS-CoV-2 at the time of analysis, allowing researchers to monitor the epidemic in a particular geographical region more effectively. Two main SARS-CoV-2 lineages are currently recognized; lineage A, defined by Wuhan/WH04/2020 and lineage B, defined by Wuhan-Hu-1 strain. Although Wuhan-Hu-1 was the first published genome from SARS-CoV-2, it is classified as lineage B. Phylogenetic analyses of SARS-CoV-2 identified sequences from lineage A to be more closely related to a bat corona virus (14), which suggest this to be the first lineage (hence A). Lineage A genomes are characterized by two unique mutations (8782C>T and 28144T>C), relative to lineage B. Lineage B, on the other hand, shares no common mutations since this lineage contains the global SARS-CoV-2 genome reference (Wuhan-Hu-1). From these lineages, sub-lineages (e.g. A·1, A·2, A·3 and so forth) are then designated, each defined by an additional set of unique mutations. For example, for sub-lineage A·1, these mutations would be; 11747C>T, 1785A>G and 18060C>T. Sub-lineages can further diversify into sub sub-lineages (e.g. A·1·1). Please refer to the schema provided in Supplementary Figure 5 for more information.

### Phylogenetic analysis

10,959 GISAID reference genomes and 20 KRISP sequences were aligned in Mafft v7·313 (FF-NS-2) followed by manual inspection and editing in the Geneious Prime software suite (Biomatters Ltd, New Zealand). We constructed a maximum likelihood (ML) tree topology in IQ-TREE (GTR+G+I, no support) (29,30). Due to the large size of the alignment and the low variability, we opted to not infer support for splits in this tree topology. In any tree topology of SARS-CoV-2 the majority splits will be poorly supported with only the major splits separating the major lineages having good support. The resulting ML tree topology was transformed into a time scaled phylogeny using TreeTime (31) with a clock rate of 8×10^−4^and rooted along the branch of Wuhan-WH04 (GISAID: hCoV19/Wuhan/WH04/2020) and Wuhan-Hu1 (Genbank: MN908947). The resulting phylogeny was viewed and annotated in FigTree and ggtree.

Based on this large phylogeny of SARS-CoV-2, we randomly down sampled the GISAID reference sequences that passed initial sequence quality checks to ∼10% of the original size. All African sequences in the GISAID subset, the 20 genotypes that were generated in this study, as well as a select few external references (e.g. Wuhan-Hu-1) were included. The resulting dataset of 1848 sequences was used in a custom build on the NextStrain analysis platform (32). To infer support for the splits in this tree topology we inferred an additional 100 bootstrap trees in IQ-Tree under the same model parameters as NextStrain. These trees were then used to infer transfer support for splits in the phylogeny (33).

### Bayesian Tree

Bayesian coalescent analyses were performed on major lineages of the NextStrain build in which KZN sequences fell. The purpose of these analyses were to: (*i*) confirm the estimated date of origin for SARS-CoV-2 as proposed in recent literature (34,35), (*ii*) infer the estimated date to the most recent common ancestor (MRCA) for major lineages and (*iii*) infer the estimated dates of viral introductions into South Africa.

Due to the dynamic lineage assignment system of pangolin, many sub sub-lineages (e.g. A·1·1 or A·1·1·1) have emerged since the start of the outbreak. In order to keep things neat and tidy we organized B lineages into B, B·1 and B·2. Due to the large number of B and B·1 lineages, we randomly down sampled these while retaining all South African genotypes. This resulted in three datasets for Bayesian coalescent inference: (B = 128, B·1 = 178 & B·2 = 69). Since none of the KZN sequences were classified as lineage A we exclude A genotypes from our Bayesian analyses.

In short, sequences were aligned in mafft v7.313 and visualized and manually edited in Geneious software (Biomatters Ltd, New Zealand) as previously described. ML-tree topologies were inferred from each alignment in IQ-TREE v1·6·9 (GTR+G+I, with transfer support values) (29,30). Resulting tree topologies were analyzed in TempEst software suite for temporal clock signal (Supplementary Figure S4). Coalescent molecular clock analyses were performed in BEAST v1·8. In short, analyses were run under a strict molecular clock assumption at a constant evolutionary rate of 8×10^−4^nucleotide substitutions per site per year and an exponential growth coalescent tree prior. The Markov Chains were run in duplicate for a total length of 100 million steps sampling every 10,000 iterations in the chains. Runs were assets in Tracer for good convergence (ESS>200) and TreeAnnotator after discarding 10% of runs as burn-in.

## Data Availability

SARS-CoV-2 genome sequences generated in this study have been deposited in the GISAID database (https://www.gisaid.org/)

## Data Availability

SARS-CoV-2 genome sequences generated in this study have been deposited in the GISAID database(https://www.gisaid.org/),under the following accession IDs: *EPI_ISL_421572, EPI_ISL_421573, EPI_ISL_421574, EPI_ISL_421575 EPI_ISL_421576 EPI_ISL_436684 EPI_ISL_436685 EPI_ISL_436686 EPI_ISL_436687*. In addition, raw short and long reads have been submitted to the Short Read Archive (SRA) and can be accessed under accession numbers ABC-XYZ.

## Acknowledgments

We wish to extend our thanks to all laboratory personnel that have worked hard to genotype SARS-CoV-2 samples and who have generously made it public via the GISAID database. Without this free data-sharing environment, this research would not have been possible. A full list of acknowledgments to contributing laboratories can be found in Supplementary Table S8.

